# Genetic downregulation of interleukin-6 signaling and arteriolosclerotic cerebral small vessel disease: a drug target Mendelian randomization analysis

**DOI:** 10.1101/2024.12.13.24318994

**Authors:** Larissa Ange Tchuisseu-Kwangoua, Murad Omarov, Alexey Shatunov, Hugh S. Markus, Joseph Kamtchum-Tatuene, Marios K. Georgakis

## Abstract

**Background:** Arteriolosclerotic cerebral small vessel disease (cSVD) is a leading cause of stroke and dementia, yet no disease-modifying therapies exist. Anti-inflammatory strategies targeting IL- 6 signaling have shown efficacy in preventing atherosclerotic cardiovascular disease, but their potential in arteriolosclerotic cSVD remains unexplored. We investigated whether genetically downregulated IL-6 signaling is associated with clinical, imaging, and pathological manifestations of arteriolosclerotic cSVD.

**Methods:** We applied two-sample Mendelian randomization (MR) using (i) 26 genetic variants near *IL6R* associated with circulating C-reactive protein (CRP) levels and (ii) rs2228145, a well-characterized *IL6R* missense variant, as proxies of IL-6 signaling downregulation. Outcomes included clinical (small-vessel stroke, MRI-defined lacunar stroke, non-lobar intracerebral hemorrhage [ICH], vascular dementia), imaging (white matter hyperintensity volume, extensive basal ganglia perivascular space, non-lobar/mixed cerebral microbleeds), and pathological (arteriolosclerosis burden in autopsy) traits of cSVD, as well as atherosclerosis traits (ultrasound-defined carotid plaque, large artery stroke) as positive controls. We used inverse-variance weighting and the Wald ratio estimator for primary analyses. MR-Egger regression, weighted median, and weighted mode estimators were used as sensitivity analyses.

**Results:** Genetically downregulated IL-6 signaling (30%-decrement in CRP via 26 *IL6R* variants) was not associated with small-vessel stroke (OR: 1.02, 95%CI: 0.95–1.10), MRI-confirmed lacunar stroke (OR: 0.95, [0.81–1.11]), non-lobar ICH (OR: 1.04, [0.72–1.50]), or vascular dementia (OR: 1.09, [0.95–1.25]). Similarly, we found no significant association with cSVD imaging biomarkers or pathology-defined arteriolosclerosis. As expected, genetically downregulated IL-6 signaling was associated with lower odds of large artery stroke (OR: 0.79, [0.74-0.84]) and carotid plaque (OR: 0.88, [0.83–0.94]). Results were consistent across sensitivity analyses and when using the rs2228145 missense variant to proxy IL-6 signaling downregulation.

**Conclusion:** Genetically proxied IL-6 signaling downregulation is not associated with clinical, imaging or pathological manifestations of arteriolosclerotic cSVD. Therefore, genetic data suggest that targeting IL-6 signaling is unlikely to prevent cSVD manifestations.

## INTRODUCTION

Cerebral small vessel disease (cSVD) encompasses a diverse set of clinical and radiological phenotypes and accounts for about 25% of ischemic strokes and nearly all cases of intracerebral hemorrhage (ICH).^1,2^ It is the leading cause of vascular dementia^3–5^ and an independent risk factor for all-cause and vascular mortality.^6,7^ MRI markers of cSVD —lacunes, white matter hyperintensities(WMH), enlarged perivascular spaces (EPVS), and cerebral microbleeds (CMBs) — are highly prevalent in the aging population appearing in up to 90% of individuals aged 65 years and older.^8–10^ Despite the significant public health burden, the mechanisms underlying cSVD remain largely elusive, which has impeded the development of effective disease-modifying therapies.^11–13^

Anti-inflammatory treatments targeting the interleukin-6 (IL-6) signaling pathway are emerging as preventive strategies for cardiovascular disease.^14^ In phase 3 trials of patients with coronary artery disease, drugs targeting upstream regulators of IL-6 signaling, such as canakinumab^15^ and colchicine^16,17^, have shown significant reductions in adverse cardiovascular events including ischemic stroke. However, it remains uncertain if these benefits extend to non-atherosclerotic vascular pathologies. Arteriolosclerosis of the deep perforating arterioles, the most common pathology finding in cSVD,^18^ is speculated to have an immune component, but the exact inflammatory mediators driving its progression remain undetermined.^19^

This uncertainty complicates patient selection for trials of anti-inflammatory treatments for secondary ischemic stroke prevention. The Colchicine for Prevention of Vascular Inflammation in Non-Cardioembolic Ischemic Stroke (CONVINCE) trial evaluated the benefit of colchicine in patients with non-cardioembolic ischemic strokes, including those attributed to cSVD.^20^ Although the study did not show a significant reduction in the primary endpoint of recurrent vascular events in the intention-to-treat analysis, a promising reduction was observed in the per-protocol analysis, excluding patients who were non-adherent to colchicine.^20^ Ongoing trials, such as CASPER or RIISC-THETIS,^21,22^ are also assessing colchicine for secondary prevention in ischemic stroke and include patients with cSVD-related infarcts.

Human genetics can offer key insights into causal disease mechanisms and inform drug development. Genetically supported drug targets are 2.6 times more likely to result in approved treatments.^23^ Variants near the gene encoding the IL-6 receptor (*IL6R*) are associated with downregulated IL-6 signaling and have served as proxies to study the potential effect of pharmacological IL-6 inhibition on various cardiovascular outcomes.^24–31^ Evidence from these genetic studies and findings from clinical^32^ and population-based studies^33^ have strengthened interest in IL-6 signaling inhibition as a preventive strategy in atherosclerotic cardiovascular disease, with several phase 3 trials currently underway (ZEUS [NCT05021835], ATHENA [NCT06200207], HERMES [NCT05636176], ARTERMIS [NCT06118281]).^34–37^ We performed a drug target Mendelian randomization to explore whether IL-6 inhibition could also be a viable option for preventing cSVD-related complications. We assessed whether genetically proxied IL-6 signaling downregulation is associated with clinical manifestations, imaging biomarkers, and pathological hallmarks of arteriolosclerotic cSVD, using atherosclerosis-related traits as positive controls.

## METHODS

This report is compliant with the Strengthening the Reporting of Observational Studies in Epidemiology using Mendelian Randomization (STROBE-MR) statement.^38^ A graphical overview of the study design is presented in **Figure 1**. The data sources used for the analyses are provided as **Supplementary Table 1**. We applied two-sample MR which uses selected genetic instruments to assess the relationship between genetically proxied IL-6 signaling and various cSVD manifestations.

**Figure 1.**
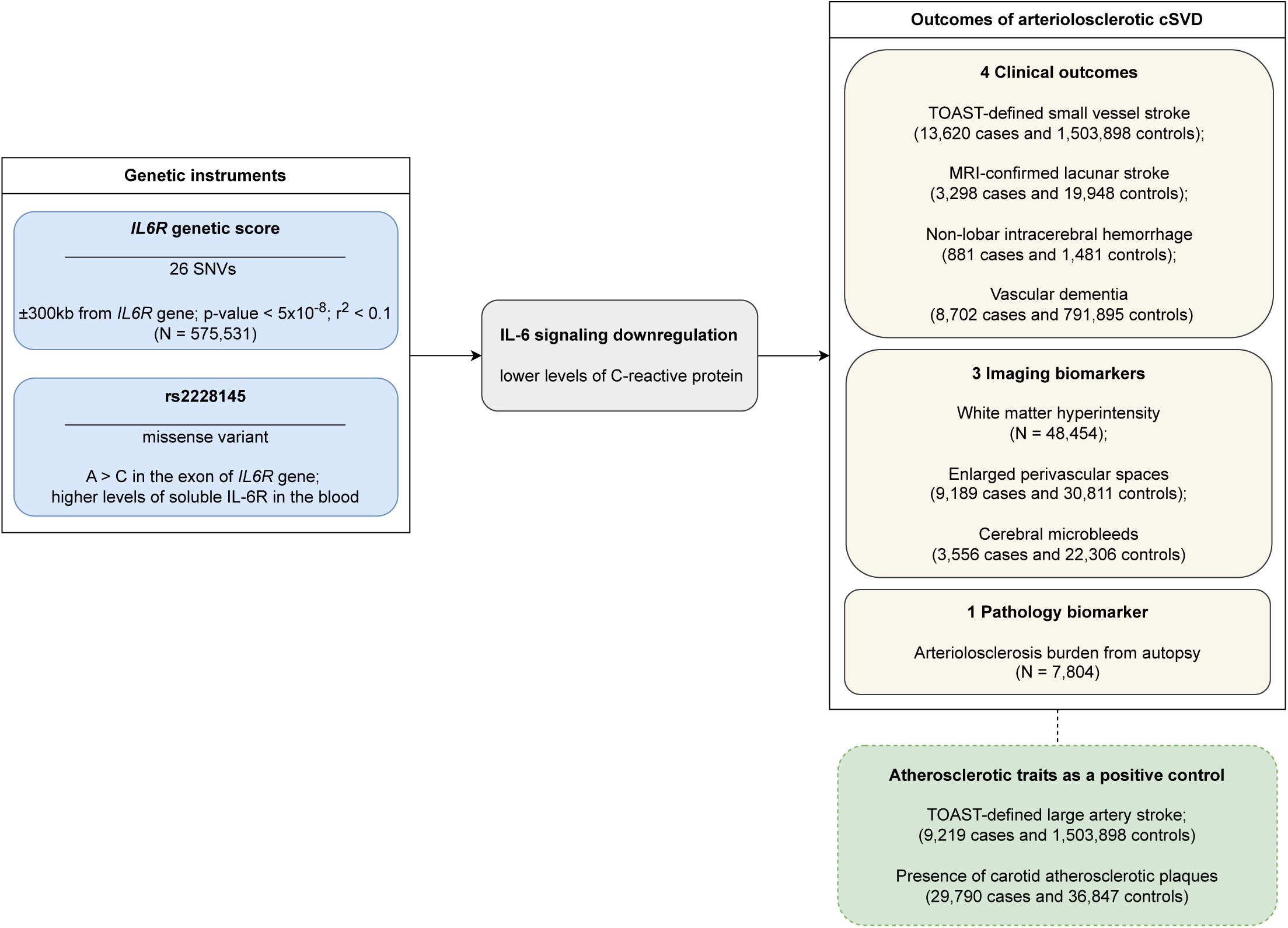
Summary of the study design and of the data sources used for the analysis. *IL6R* – Interleukin-6 receptor; SNV – Single nucleotide variant; IL-6 – Interleukin-6; cSVD – Cerebral small vessel disease

### Genetic instruments

We used two sets of genetic instruments to proxy IL-6 signaling downregulation. First, according to an approach we previously developed,^26,27,30,39^ a list of 26 genetic variants, were selected based on (i) their location within a window including the *IL6R* gene ±300 kb; (ii) their association with circulating C-reactive protein (CRP) levels at a p-value < 5x10^-8^ in a GWAS meta-analysis of 575,531 individuals of European ancestry from the Cohorts for Heart and Aging Research in Genomic Epidemiology (CHARGE) consortium and the UK Biobank;^40^ and (iii) their lack of strong correlation between them due to linkage disequilibrium (clumped at r^2^ < 0.1). Second, we used the missense single nucleotide polymorphism (SNP) rs2228145, for which there is strong evidence for a functional effect leading to downregulation of IL-6 signaling and a significant reduction in circulating levels of CRP.^24^ This SNP is located within the *IL6R* gene and leads to a change from adenine (A) to cytosine (C) at position 1073 in the 9^th^ exon of the gene resulting in a Asp358 → Ala358 change in the IL-6R protein. The Asp358Ala substitution leads to an increase in the shedding of the membrane-bound IL-6R (mIL-6R), thus decreasing the availability of mIL- 6R and increasing the production of circulating soluble IL-6R (sIL-6R).^41,42^

For both instruments we calculated the F-statistics of the individual variants, as well as the cumulative variance of CRP explained by all variants in an additive model. Detailed information on the instruments is provided in **Supplementary Table 2**.

### Analysis of individual-level UK Biobank data

We used data from the UK Biobank to explore the effects of both instruments on circulating CRP levels. UK Biobank is a UK-wide prospective cohort of 503,317 adults aged 39-73, recruited between 2006 and 2010. The design and rationale of the study have been described previously.^43^ Eligible participants were registered with the National Health Service in England, Scotland or Wales, and lived close to one of the 22 assessment centers in these regions. Details on demographics, lifestyle and medical history were collected during visit to the assessment centers, along with anthropometric data and biological (blood, urine and saliva) samples collection by trained personnel. Participants gave their written informed consent to be followed up through a national record linkage.

We analyzed individual-level data from 487,067 participants with available genotypic data. Initially, genotyping was performed for 488,377 participants, with 49,979 genotyped using the UK BiLEVE array and 438,398 using the UK Biobank Axiom array. Following standard quality control procedures, individuals with sex-mismatch (derived by comparing genetic sex and reported sex) or individuals with sex chromosome aneuploidy were excluded from the analysis, leading to a total of 487,067 individuals.^44^ Pre-imputation quality control, phasing and imputation are described elsewhere.^44^ In brief, prior to phasing, multiallelic SNPs or those with minor allele frequency ≤1% were removed. Phasing of genotype data was performed using a modified version of the SHAPEIT2 algorithm.^45^ Genotype imputation to a reference set combining the UK10K haplotype and HRC reference panels was performed using IMPUTE2 algorithms.^46^

For the first instrument, we used the clumping and thresholding method to construct a genetic risk score based on the 26 *IL6R* CRP-increasing genetic variants and explored differences in CRP between the top and bottom percentile of the distribution.^47^ The score was calculated with *plink v2.00a*.^48^ For the rs2228145, we calculated differences in CRP between carriers of the A and C alleles of variant. We explored the effects of the two genetic instruments on ln-transformed CRP levels in linear regression models adjusted for age, sex, and the first 10 principal components of the population structure – we also ran univariable linear regression analyses to calculate the proportion of CRP explained by the two instruments. We scaled both instruments to correspond to a reduction in CRP levels that reflects the natural variation observed across carriers of the genetic variants (30% reduction in median CRP levels for individuals in the bottom vs. top percentile of the distribution of the 26-variant genetic score and 18% reduction in median CRP levels for CC vs. AA allele carriers).

### Outcomes

We use clinical outcomes of cSVD, namely lacunar stroke, non-lobar ICH, and vascular dementia. In addition, we analyzed associations with MRI markers of cSVD. This allows accurate phenotyping of the features of cSVD but provides information on the more chronic features of cSVD, rather than risk factors for acute infarction. We also studied associations with pathology-confirmed arteriolosclerosis. We used publicly available summary statistics from genome-wide association studies (GWAS) to extract the effects of the selected genetic variants on the outcomes of interest (**Supplementary Table 1**).

### Clinical manifestations attributed to arteriolosclerotic cerebral small vessel disease

i. Small vessel stroke defined as a clinical syndrome of lacunar stroke without evidence of alternative etiologies in diagnostic workup, and no evidence of a new cortical or large subcortical [≥ 15 mm] infarct on CT or MRI as per TOAST criteria.^49^ We used data from the cross-ancestry GWAS meta-analysis of the GIGASTROKE study (13,620 cases and 1,503,898 controls; 67% European, 25% East Asian, 4% African, 3% South Asian, 1% Hispanic)^50^;
ii. MRI-confirmed lacunar stroke. Small vessel stroke in the GIGASTROKE database has been subtyped based largely on a diagnosis of a clinical lacunar syndrome combined with CT brain imaging. However, CT imaging may not show lacunar infarcts, particularly in the first 24 hours. MRI provides a more accurate diagnosis of lacunar infarction, since as many as 50% of all patients classified as small vessel stroke based on CT imaging do not have cSVD on MRI .^51^ Therefore, we also studied a population with MRI-confirmed lacunar stroke defined as a clinical lacunar stroke syndrome stroke with an anatomically corresponding lacunar infarct of ≤ 15 mm in diameter. We used data from a study of 3,298 cases and 19,948 controls (only European ancestry), which included data from a published study (2,612 cases)^52^ and 686 newly genotyped samples from the UK DNA Lacunar Stroke studies 1 and 2.
iii. Primary spontaneous intracerebral hemorrhage (ICH) originating in non-lobar brain locations (thalamus, internal capsule, basal ganglia, deep periventricular white matter, cerebellum, or brain stem) as confirmed with neuroimaging. We used data from a GWAS meta-analysis of 6 case-controls studies of European ancestry only.^53^ Only non-lobar ICH cases (corresponding to 881 cases and 1,481 controls) were analyzed, as they are more likely to be etiologically related to arteriolosclerosis, as opposed to lobar ICH cases that are more commonly linked to cerebral amyloid angiopathy.^54^
iv. Vascular dementia defined according to clinical criteria (DSM III to V, ICD-9 or 10, NINDS-AIREN, and ADDTC). We used data from a GWAS meta-analysis of 21 cohorts and consortia performed by the Mega Vascular Cognitive Impairment and Dementia MEGAVCID Consortium (8,702 cases and 753,695 controls; 97% European, 1% African, 1% East Asian, <1% Hispanic).^55^ Only the European-specific GWAS was used in the analysis made of 3,892 cases and 466,606 controls. cSVD is considered one of the major contributors to vascular dementia.^3–5^

### Imaging biomarkers of arteriolosclerotic cerebral small vessel disease

i. White matter hyperintensity (WMH) volume on brain MRI images (T2 weighted or FLAIR sequences) quantified with fully automated or semi-automated software. We extracted data from a GWAS meta-analysis of predominantly European ancestry participants of the UK Biobank and 23 CHARGE cohorts (N = 48,454, 95% European and 5% African-American).^56^ WMH volume was inverse-normal transformed across the cohorts — association estimates may be interpreted in standard deviation units of WMH volume.
ii. Enlarged perivascular spaces (EPVS) defined as fluid-filled spaces with a signal identical to that of CSF with a maximum diameter smaller than 3 mm, no hyperintense rim on T2-weighted or FLAIR sequences, and located in areas supplied by perforating arteries. We obtained data from a GWAS meta-analysis of 18 population-based studies (UK Biobank, cohorts of the CHARGE Consortium and the BRIDGET Initiative) including 40,095 stroke-free participants (97% European, 2% Hispanic, <1% East Asian, <1% African-American).^57^ Across the provided locations (white matter, basal ganglia, hippocampus), we focused on EPVS in basal ganglia, as these lesions are more likely to be related to arteriolosclerosis.^58^ As different scales were used to quantify EPVS across studies, the trait was analyzed as a binary variable with logistic regression (extensive vs. non-extensive EPVS; 9,189 cases and 30,811 controls) with a cohort-specific threshold closest to the top quartile of the semiquantitative scale distribution within each cohort.57
iii. Cerebral microbleeds (CMB) recognized as small hypointense lesions on susceptibility-weighted imaging (SWI) sequences or, to a lesser extent, on T2*-weighted gradient echo sequences. Data on CMB were extracted from a GWAS meta-analysis undertaken in population-based cohorts of the CHARGE consortium and the UK Biobank, totaling 25,862 individuals of all ancestries (3,556 cases with microbleeds, of which 2,179 strictly lobar and 1,293 deep, infratentorial, or mixed, and 22,306 controls; 97% European, 2% African American, <1% Malay, <1% Chinese).^59^ We focused our analyses on deep or infratentorial CMBs and mixed CMBs, as they are more likely to be etiologically related to arteriolosclerosis, as opposed to strictly lobar CMBs that are typically linked to cerebral amyloid angiopathy.^54^

### Pathology burden of arteriolosclerosis

To explore associations with pathology-confirmed arteriolosclerosis, we extracted data from a GWAS meta-analysis of data from the US National Alzheimer’s Coordinating Center (NACC) neuropathology study, the Religious Orders Study and Memory and Aging Project (ROSMAP), and the Adult Changes in Thought (ACT) study totaling 7,804 autopsied participants (all European ancestry).^60^ Arteriolosclerosis burden was analyzed as an ordinal variable ranging from 0 to 3 (0 = “none”, 1 = “mild”, 2 = “moderate”, 3 = “severe”) according to qualitative assessment of a neuropathologist.^61^

### Atherosclerosis-related traits as positive controls

Given the established previous associations of genetic proxies of IL-6 signaling downregulation with atherosclerotic cardiovascular disease^24–31^, we analyzed cerebrovascular atherosclerosis traits as positive controls:

i. TOAST-defined large artery atherosclerotic stroke defined as an ischemic stroke with evidence of a ≥ 50% stenosis in a supplying artery. We used data from the trans-ancestry GIGASTROKE study (9,219 cases and 1,503,898 controls; 67% European, 25% East Asian, 4% African, 3% South Asian, 1% Hispanic);^50^
ii. Presence of carotid atherosclerotic plaques on carotid ultrasound defined as focal wall structure protruding into the arterial lumen of the distal common or proximal internal carotid artery by ≥ 0.5 mm or ≥50% of the surrounding intima-media thickness or with an overall thickness of > 1.5 mm as measured from the media-adventitia interface to the intima-lumen interface.^62^ We used data from a GWAS meta-analysis of the UK Biobank and the CHARGE cohorts (29,790 cases and 36,847 controls; all European ancestry).^63^

### Statistical analysis

We performed two-sample Mendelian randomization (MR) to explore the associations between genetically proxied downregulation of IL-6 signaling and the cSVD-related outcomes. Mendelian Randomization analyses were performed using the TwoSampleMR package in R Studio v3.5.1.For the 26-variant instrument, we used fixed-effects inverse variance-weighted (IVW) MR as our primary analytical approach.^64^ The Cochran Q test was used to assess heterogeneity between the variants as a metric of pleiotropy (statistical significance set at p<0.05).^65^ To ensure the robustness of the results against the assumptions of IVW MR, three other MR sensitivity methods were used:

MR-Egger, weighted mode, and weighted median. MR Egger regression assumes that genetic instruments are uncorrelated with any pleiotropic effect of the instrument on the outcome.^66^ The intercept of MR Egger regression was used as a measure of unbalanced pleiotropy (p<0.05 indicated significance). The weighted mode assumes that the most common effect is consistent with the true causal effect.^67^ The weighted median assumes that at least 50% of the weight comes from valid SNPs. For the rs2228145 variant, we applied the Wald ratio method.^68^

The derived odds ratios (OR) correspond to a 30% or 18% decrement in CRP levels for the 26- variant instrument and the rs2228145 variant, respectively. All p-values are two-sided. As we analyzed 8 main outcomes (4 clinical, 3 radiological, 1 pathological) our significance threshold was set at p < 0.00625 to account for multiple testing using the Bonferroni approach. A p < 0.05 indicates nominal significance. For clinical outcomes, we performed power calculations estimating the OR range we were sufficiently powered (1-β > 0.8) to detect at a nominal significance level (α = 0.05).

### Ethics

The UK Biobank has obtained approval from the Northwest Multi-Center Research Ethics Committee (REC reference for UK Biobank is 11/NW/0382). All participants have provided written informed consent. Data for these analyses were accessed under application number 151281. DNA Lacunar 2 was approved by East of England Research Ethics Committee (16/EE/0201) and all participants provided signed informed consent. The publicly available GWAS summary statistics were generated by studies that had obtained ethical approval and participant consent for analyses and distribution of summary-level data, as described in the original publications.

### Data availability

The genetic variants used as instruments in this analysis along with their weights are provided in **Supplementary Table 2**. The GWAS summary datasets for CRP used to weigh the instruments are downloadable from the GWAS catalog under accession number GCST90029070. Summary statistics for small vessel stroke, extended basal ganglia perivascular space and large artery stroke are available from the GWAS Catalog under accession numbers GCST90104537, GCST90244154 and GCST90104542. GWAS summary data from the CHARGE Consortium on WMH and non- lobar ICH can be obtained from the Database of Genotypes and Phenotypes (dbGaP) under accession numbers phs002227.v1.p1 and phs000416.v1.p1. GWAS summary statistics for CMBs are available through the <u>Dataset Inspector Cerebrovascular Disease Knowledge Portal</u>. GWAS summary statistics for ultrasound-defined carotid plaque are available from the GWAS Catalog under the accession number GCST90454353. GWAS summary statistics for vascular dementia, MRI-confirmed lacunar stroke, and pathology-defined arteriolosclerosis burden were provided upon request to the corresponding authors of the articles describing the datasets. UK Biobank data are available upon submission of an application to the UK Biobank.

## RESULTS

### Genetic instruments for IL-6 signaling downregulation

A total of 487,067 participants from the UK Biobank (54.1% female, median age 59 years) had available genetic data and were included in this analysis (**Table 1**). A genetic score consisting of the 26 variants included in our main genetic instrument was strongly associated with CRP, explaining 0.45% of its variance in a linear regression model. Individuals in the top percentile of the distribution of the score had 30.3% higher CRP levels on average, when compared with individuals in the bottom percentile (1.52 vs. 1.06 mg/dL, **Table 2**). Among the study participants, there were 230,868 (47.4%) heterozygote and 79,949 (16.4%) homozygote carriers of the C allele of the rs2228145 variant. When compared with homozygote carriers of the A allele, the median serum CRP levels were 8.8% and 18.2% lower in participants with AC and CC genotypes, respectively (median CRP levels: 1.48, 1.35, 1.21 mg/dL for AA, AC, and CC genotypes, respectively, **Table 2**).

**Table 1.**
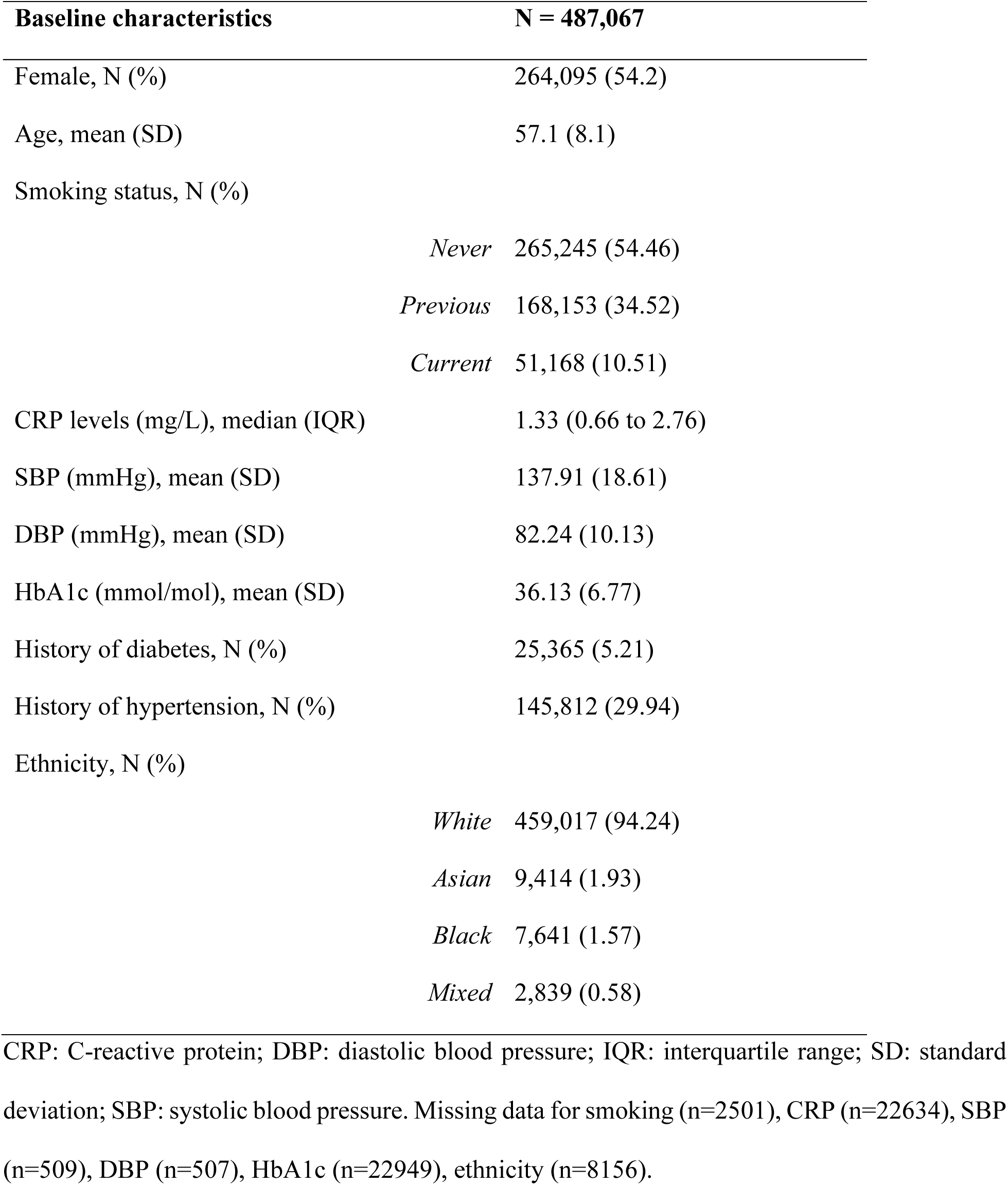
Baseline characteristics of the UK Biobank participants with available genetic data.

**Table 2.**
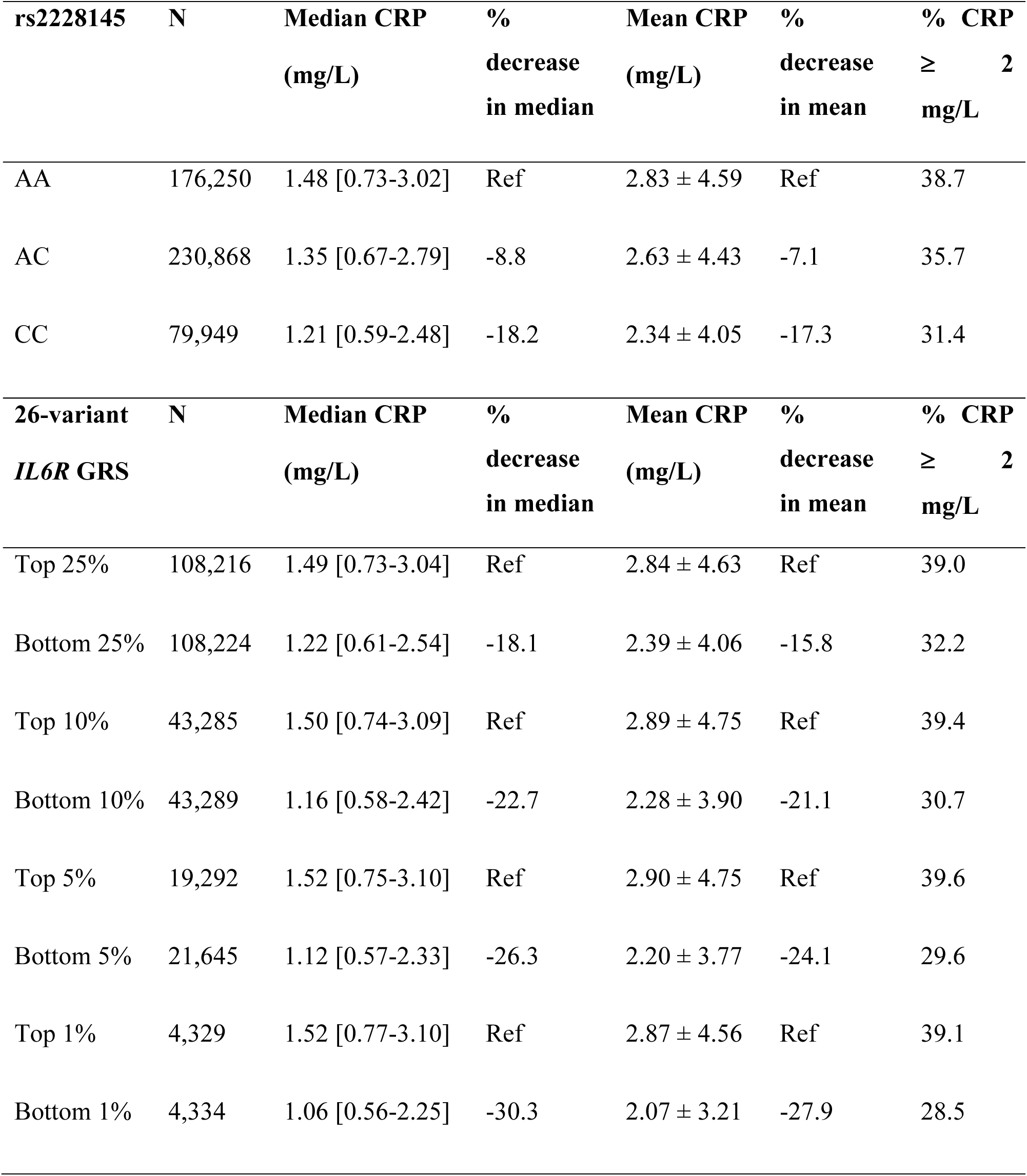
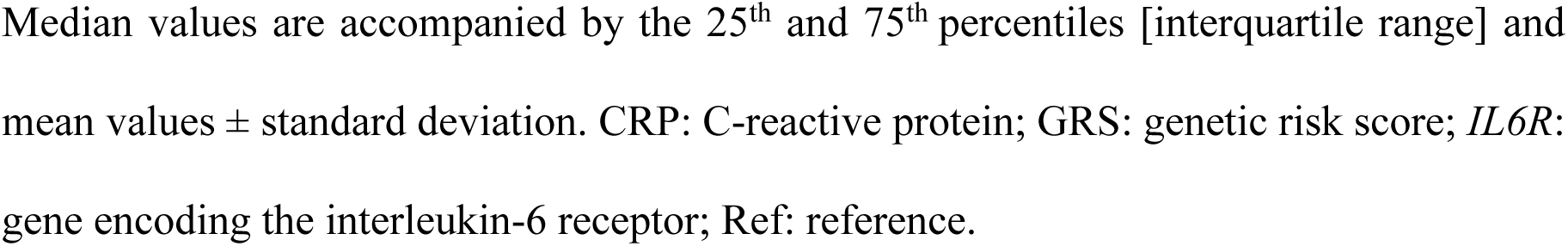
C-reactive protein levels across the distribution of the genetic instruments for IL-6 signaling downregulation in the UK Biobank (N=487,067).

| **rs2228145** | **N** | **Median CRP**<br>**(mg/L)** | **%**<br>**decrease**<br>**in median** | **Mean CRP**<br>**(mg/L)** | **%**<br>**decrease**<br>**in mean** | **% CRP**<br>**≥ 2**<br>**mg/L** |
| --- | --- | --- | --- | --- | --- | --- |
| AA | 176,250 | 1.48 [0.73-3.02] | Ref | 2.83 ± 4.59 | Ref | 38.7 |
| AC | 230,868 | 1.35 [0.67-2.79] | -8.8 | 2.63 ± 4.43 | -7.1 | 35.7 |
| CC | 79,949 | 1.21 [0.59-2.48] | -18.2 | 2.34 ± 4.05 | -17.3 | 31.4 |
| **26-variant**<br>**IL6R GRS** | **N** | **Median CRP**<br>**(mg/L)** | **%**<br>**decrease**<br>**in median** | **Mean CRP**<br>**(mg/L)** | **%**<br>**decrease**<br>**in mean** | **% CRP**<br>**≥ 2**<br>**mg/L** |
| Top 25% | 108,216 | 1.49 [0.73-3.04] | Ref | 2.84 ± 4.63 | Ref | 39.0 |
| Bottom 25% | 108,224 | 1.22 [0.61-2.54] | -18.1 | 2.39 ± 4.06 | -15.8 | 32.2 |
| Top 10% | 43,285 | 1.50 [0.74-3.09] | Ref | 2.89 ± 4.75 | Ref | 39.4 |
| Bottom 10% | 43,289 | 1.16 [0.58-2.42] | -22.7 | 2.28 ± 3.90 | -21.1 | 30.7 |
| Top 5% | 19,292 | 1.52 [0.75-3.10] | Ref | 2.90 ± 4.75 | Ref | 39.6 |
| Bottom 5% | 21,645 | 1.12 [0.57-2.33] | -26.3 | 2.20 ± 3.77 | -24.1 | 29.6 |
| Top 1% | 4,329 | 1.52 [0.77-3.10] | Ref | 2.87 ± 4.56 | Ref | 39.1 |
| Bottom 1% | 4,334 | 1.06 [0.56-2.25] | -30.3 | 2.07 ± 3.21 | -27.9 | 28.5 |

Median values are accompanied by the 25<sup>th</sup> and 75<sup>th</sup> percentiles [interquartile range] and mean values ± standard deviation. CRP: C-reactive protein; GRS: genetic risk score; *IL6R*: gene encoding the interleukin-6 receptor; Ref: reference.

### Genetically downregulated IL-6 signaling and clinical arteriolosclerotic cSVD outcomes

The F-statistics for the 26 selected variants ranged from 31 to 2033, thus indicating sufficient instrument strength. In line with previous studies,^30,31,63,69^ genetically downregulated IL-6 signaling proxied by the 26-variant genetic instrument was associated with lower odds of large artery atherosclerotic stroke (OR per 30% decrement in CRP levels: 0.79, 95%CI: 0.72–0.87, p = 4×10^-6^) in the GIGASTROKE study (**Figure 2A**). However, we found no evidence that genetically proxied downregulation of IL-6 signaling is associated with cSVD-related clinical outcomes including TOAST-defined small vessel stroke (OR: 1.02, 95%CI: 0.95–1.10, p = 0.54), MRI-confirmed lacunar stroke (OR: 0.95, 95%CI: 0.81–1.11, p = 0.50), non-lobar intracerebral haemorrhage (OR: 1.04, 95%CI: 0.72–1.50, p = 0.84), or vascular dementia (OR: 1.09, 95%CI: 0.95–1.25, p = 0.221, **Figure 2A)**. For all four outcomes, the ORs were significantly larger than the OR for large artery stroke, as evidenced by significant heterogeneity estimates (all p < 0.05, **Figure 2A**). *Post hoc* power calculations for the 26-variant instrument suggested that the datasets used for TOAST-defined small vessel stroke, MRI-confirmed lacunar stroke, and vascular dementia offered sufficient statistical power to detect associations similar to the one detected for large artery stroke (**Supplementary Table 3**). With the exception of vascular dementia (p-value of Q heterogeneity = 0.0067), there was no evidence of between-variant heterogeneity for any of the outcomes. The alternative MR methods (MR Egger regression, weighted mode, and weighted median estimator) for the 26-variant instrument showed similar effects as the IVW analysis (**Supplementary Table 4**).

**Figure 2.**
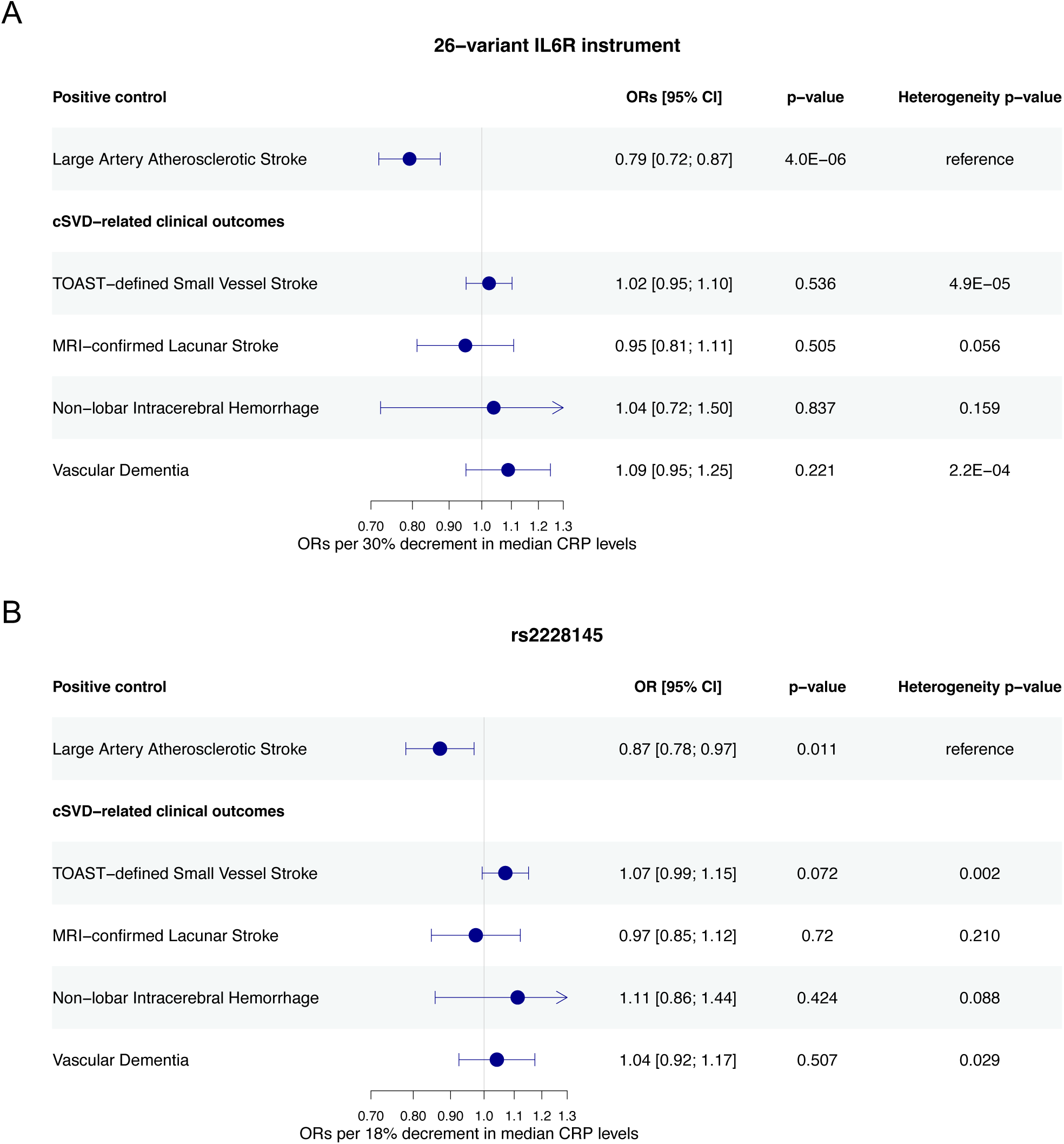
Genetically proxied IL-6 signaling downregulation and arteriolosclerotic cSVD- related clinical outcomes. Results derived from (**A**) a fixed-effects inverse-variance weighted Mendelian randomization analysis for a genetic instrument composed of 26 CRP-lowering variants in the *IL6R* locus and (**B**) the Wald ratio method for the genetic instrument composed of the single rs2228145 variant. Heterogeneity p-values calculated using the Cochran’s Q statistic represent comparisons between the OR for each outcome and OR for large artery atherosclerotic stroke (positive control). *IL6R*: Interleukin-6 receptor; cSVD – Cerebral small vessel disease; CRP – C-reactive protein.

**Figure 3.**
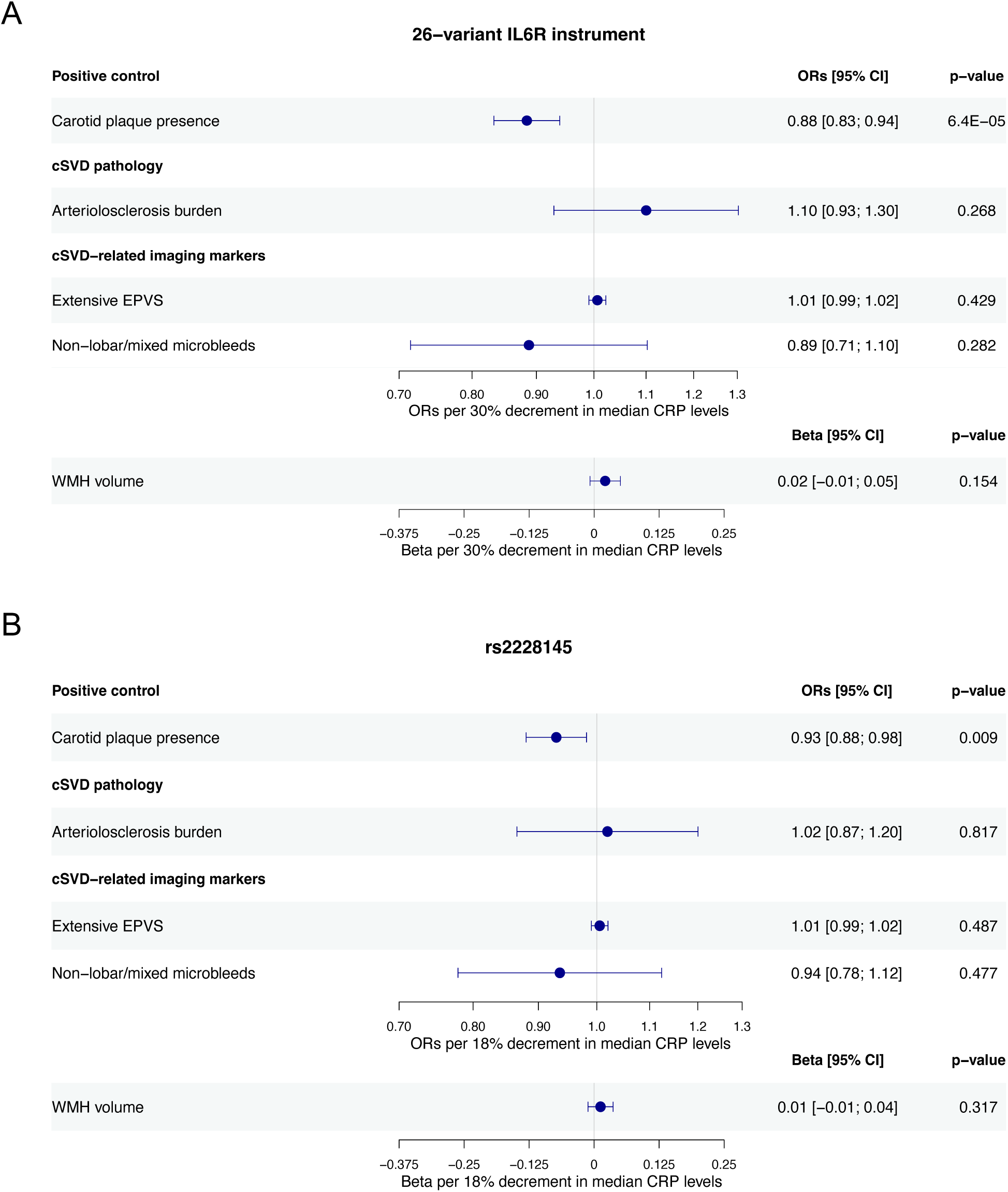
Genetically proxied IL-6 signaling downregulation and arteriolosclerotic cSVD- related imaging and pathology outcomes. Results derived from (**A**) a fixed-effects inverse-variance weighted Mendelian randomization analysis for a genetic instrument composed of 26 CRP-lowering variants in the *IL6R* locus and (**B**) the Wald ratio method for the genetic instrument composed of the single rs2228145 variant. CRP – C-reactive protein; cSVD – Cerebral small vessel disease; EPVS – Enlarged perivascular spaces; WMH – White matter hyperintensities

The analysis for the missense variant rs2228145 revealed similar associations, albeit, as expected, of wider confidence intervals due to lower statistical power (**Figure 2B**). Specifically, there was a nominal association between genetically proxied IL-6 signaling downregulation and lower odds of large artery stroke (OR per 18% decrement in CRP levels: 0.87, 95%CI: 0.78–0.97, p = 0.01), but no evidence of nominal or significant association with TOAST-defined small vessel stroke, MRI-confirmed lacunar stroke, non-lobar intracerebral haemorrhage, or vascular dementia.

### Genetically downregulated IL-6 signaling and imaging and pathological arteriolosclerotic cSVD traits

After correcting for multiple comparisons, genetically proxied downregulation of IL-6 signaling through either of the two instruments in the *IL6R* locus was not associated with arteriolosclerotic cSVD-related imaging biomarkers (WMH volume, deep/mixed CMBs, extensive basal ganglia EPVS) or arteriolosclerosis burden assessed by autopsy (**Figure 4**). In contrast, there was a significant association between genetically downregulated IL-6 signaling and lower odds of a carotid plaque on ultrasound (OR per 30% decrement in CRP levels: 0.88, 95%CI: 0.83–0.94, p = 6×10^-5^). These results were consistent across different MR methods (**Supplementary Table 5**).

## DISCUSSION

In this integrative analysis of genomic, clinical, imaging, and pathological data, we found no genetic support for IL-6 signaling as a viable therapeutic target for lowering the burden of arteriolosclerotic cSVD. Specifically, IL-6 signaling activity, proxied by CRP-lowering variants near *IL6R*, was not associated with clinical outcomes related to arteriolosclerotic cSVD, including small vessel stroke, imaging-confirmed lacunar stroke, non-lobar intracerebral hemorrhage, or vascular dementia. Furthermore, there was no evidence linking genetically proxied IL-6 signaling activity to imaging markers of cSVD — WMH, CMB, and EPVS — or arteriolosclerosis pathology burden in the deep perforating arterioles of the brain in autopsied individuals.

These findings have important implications for ongoing and planned clinical trials testing anti-inflammatory treatments for secondary ischemic stroke prevention. Most of the proposed interventions are expected to cause direct or indirect inhibition of the NLRP3/IL-1β/IL-6 axis, as there is strong evidence for its involvement in atherosclerosis. Inclusion of patients with lacunar stroke or cSVD in such trials has been a matter of controversy. For example, the CONVINCE trial tested colchicine in patients with a recent history of non-cardioembolic stroke, including patients with cSVD-related stroke,^20^ but did not show a significant effect on its primary endpoint of recurrent vascular events. However, a significant reduction in risk was observed for patients with advanced atherosclerosis, as captured by a >50% carotid stenosis or a history of coronary artery disease. *Post hoc* analyses will clarify whether the inclusion of patients with lacunar stroke diluted the effect of colchicine in CONVINCE.^20^ Ongoing trials vary in their inclusion criteria: CASPER, for example includes ischemic strokes of any etiology, as long as CRP is elevated, whereas RIISC- THETIS requires a ≥ 30% stenosis in a brain-supplying artery.^21,22^

Our data for lacunar stroke appear to contradict results from earlier analyses.^30,69^ However, this discrepancy may be related to the inclusion of non-cSVD-related lacunar strokes in earlier GWAS studies of small vessel stroke. Non-MRI-defined small vessel strokes are identified by the absence of a larger cortical or subcortical infarct on CT, without evidence of a 50% stenosis in brain-supplying arteries or a cardioembolic source.^49^ This definition may include smaller infarcts associated with branch atherosclerosis disease affecting small perforating arterioles or other symptomatic non-stenotic atherosclerotic lesions.^70–72^ However, after restricting the analysis to MRI-confirmed lacunar infarcts more likely to be caused by arteriolosclerotic cSVD, we found no evidence of an effect of genetically proxied IL-6 signaling downregulation. The enrichment of the latest small vessel stroke GWAS dataset (GIGASTROKE) with MRI-confirmed lacunar strokes may also explain the negative findings for small vessel stroke in our analysis. Combined with the strong association of genetically proxied IL-6 signaling with large artery atherosclerotic stroke, these results suggest that the associations observed in earlier datasets may have been driven by a substantial proportion of traditionally-defined small vessel strokes caused by atherosclerosis.

This study has limitations. First, an inherent weakness of Mendelian randomization is its assessment of the lifelong effect of genetic perturbation of a drug target, which may not align with the outcomes of a short-term pharmacological target modulation. Only data from clinical trials testing inhibitors of IL-6 signaling can directly address this question. Second, we relied on genomic data from primarily European populations, potentially limiting the transferability of our analyses to other ancestries. Third, although we focused on genetic variants near *IL6R* to reduce pleiotropic effects, we cannot fully exclude the potential influence of SNPs impacting neighboring genes. However, the generally consistent results of an analysis focusing on a well-characterized variant within the exonic sequence of *IL6R* argue against this possibility. Fourth, despite utilizing the largest available datasets for cSVD-related outcomes, several of these GWAS analyses remain underpowered, meaning that effects of smaller magnitude cannot be entirely ruled out based on our analysis.

In conclusion, we found no evidence that genetically proxied IL-6 signaling is associated with clinical, imaging or pathology-defined outcomes related to arteriolosclerotic cSVD. Therefore, genetic data do not support the hypothesis that pharmacological therapies targeting IL-6 signaling would prevent cSVD-related adverse outcomes.

## Author contributions

MKG and JK-T conceived the study and supervised the research. LAT-K, MO and MKG performed the analyses. MKG, LAT-K, MO, and JK-T wrote and edited the manuscript. All authors provided critical revisions and approved the final version.

## Supplementary material

Supplementary Tables ST1-6

## Non-standard Abbreviations and Acronyms

ACT: Adult Changes in Thought
ADDTC: Alzheimer’s Disease Diagnostic and Treatment Center
CASPER: Colchicine After Stroke Event to Prevent Event Recurrence
CHARGE: Cohorts for Heart and Aging Research in Genomic Epidemiology
CMB: Cerebral Microbleeds
cSVD: Cerebral Small Vessel Disease CRP C Reactive Protein
CI: Confidence Interval
CONVINCE: Colchicine for Prevention of Vascular Inflammation in Non-Cardioembolic Ischemic Stroke
CT: Computed Tomography
dbGaP: Database of Genotypes and Phenotypes
DSM: Diagnostic and Statistical Manual of Mental Disorders)
EPVS: Enlarged Perivascular Spaces
GWAS: Genome-Wide Association Studies
HRC: Haplotype Reference Consortium
ICD: International Classification of Diseases ICH Intracerebral Hemorrhage
IL- 6: Interleukin-6
*IL6R*: Interleukin-6 Receptor
IVW: Inverse Variance Weighting
LAS: Large Artery Stroke
MAF: Minor Allele Frequency
MEGAVCID: Mega Vascular Cognitive Impairment and Dementia
MR: Mendelian Randomization
MRI: Magnetic Resonance Imaging
NACC: US National Alzheimer’s Coordinating Center
NCT: National Clinical Trial identifier number
NINDS-AIREN: National Institute of Neurological Disorders and Stroke - Association Internationale pour la Recherche et l’Enseignement en Neurosciences
OR: Odd Ratio
RIISC-THETIS: Reducing Inflammation in Ischemic Stroke with Colchicine, and Ticagrelor in High-risk patients-Extended Treatment in Ischemic Stroke
ROSMAP: Religious Orders Study and Memory and Aging Project SNPs Single Nucleotide Polymorphisms
STROBE-MR: Strengthening the Reporting of Observational Studies in Epidemiology using Mendelian Randomization
TOAST: The Trial of Org 10172 in Acute Stroke Treatment
UK: United Kingdom
WMH: White Matter Hyperintensity

